# How important are parents in the development of child anxiety and depression? A genomic analysis of parent-offspring trios in the Norwegian Mother Father and Child Cohort Study (MoBa)

**DOI:** 10.1101/2020.04.14.20064782

**Authors:** Rosa Cheesman, Espen Moen Eilertsen, Yasmin I. Ahmadzadeh, Line C. Gjerde, Laurie J. Hannigan, Alexandra Havdahl, Alexander I. Young, Thalia C. Eley, Pål R. Njølstad, Per Magnus, Ole A. Andreassen, Eivind Ystrom, Tom A. McAdams

## Abstract

**Background:** Many studies detect associations between parent behaviour and child symptoms of anxiety and depression. However, most do not account for shared genetic risk. Quantitative genetic designs provide a means of controlling for shared genes, but rely on observed putative exposure variables, and require data from highly specific family structures.

**Methods:** The intergenerational genomic method, Relatedness Disequilibrium Regression (RDR), indexes environmental effects of parents on child traits using measured genotypes. RDR estimates how much the parent genome influences the child indirectly via the environment, over and above effects of genes acting directly in the child. This ‘genetic nurture’ effect is agnostic to parent phenotype and captures unmeasured heritable parent behaviours. We applied RDR in a sample of 11,598 parent-offspring trios from the Norwegian Mother, Father and Child Cohort Study (MoBa) to estimate parental genetic nurture separately from direct child genetic effects on anxiety and depression symptoms at age 8. We also tested for mediation of genetic nurture via maternal emotional symptoms. Results were compared to a complementary non-genomic pedigree model.

**Results:** Parental genetic nurture significantly influenced depression symptoms at age 8, explaining 14% of the phenotypic variance. Subsequent analyses suggested that maternal anxiety and depression partially mediated the parental genetic nurture effect. The genetic nurture effect was mirrored by the finding of shared family environmental influence in our complementary pedigree model. In contrast, variance in anxiety symptoms was not significantly influenced by common genetic variation in children or parents, despite a moderate pedigree heritability.

**Conclusions:** Genomic methods like RDR represent new opportunities for genetically sensitive family research in humans, which until now has been largely confined to adoption, twin and other pedigree designs. Our results are relevant to debates about the role of parents in the development of emotional problems in children, and possibly where to intervene to reduce problems.

## Introduction

Estimating how much parental behaviour influences children’s symptoms of anxiety and depression is important for understanding causes and designing interventions, but this is challenging. It cannot be assumed that associations between parental factors, such as control and hostility, and child outcomes, such as anxiety and depression (Yap et al. 2014; McLeod et al. 2007a; McLeod et al. 2007b), represent modifiable environmental effects. Genes shared by parents and children may lead to spurious or inflated intergenerational associations. For instance, evidence suggests that the link between parental divorce and emotional problems in adult offspring is due to a common genetic component influencing parent and child traits, rather than an environmental effect of divorce (D’Onofrio et al. 2007).

Twin and pedigree studies can separate genetic and environmental influences, and results suggest that environmental effects of parents on child traits, including anxiety and depression, are weak (Plomin et al. 2016). Specifically, the absence of shared environmental influence points against effects of the parental behaviours that are useful modifiable exposures – those with systematic effects causing familial aggregation. Child anxiety symptoms are moderately genetically influenced, and the remaining variation is non-shared environmental in origin (Waszczuk et al. 2014). For depression, shared environmental influences are more commonly detected, but are transient and decline across development (Hannigan et al. 2017). However, twin and pedigree designs alone have limited utility for estimating parent effects. Shared environmental influence could be masked by interactions between genetic and shared environmental effects, which load onto the genetic component (Rijsdijk & Sham 2002). Moreover, shared environment estimates include effects of other sources of sibling resemblance (e.g. common friends), not only parents. Crucially, in the classical twin model, genes and environments are assumed to be uncorrelated, which can lead to biased estimates of shared environmental variance (Coventry & Keller, 2005; Neiderhiser et al. 2004).

Adoption and children of twins (CoT) designs provide stronger tests for environmental effects of parents on children than twin studies do, since they combine data on parents and children to correct for shared genes. Intergenerational genetically sensitive research on child anxiety is scarce, but two studies suggest that parent-child associations for anxiety remain after controlling for genetic relatedness (Ahmadzadeh et al. 2019; Eley et al. 2015). The evidence base is larger for child depression. Numerous adoption and CoT studies indicate an environmental effect of parent depression above genetic confounding (Kendler et al. 2018; Silberg, Maes, & Eaves, 2010; Singh et al. 2011; Natsuaki et al. 2014). Harsh physical punishment and marital instability are also plausible environmental factors affecting child depression, which survive correction for genetic relatedness (D’Onofrio et al. 2005; Lynch et al. 2006). Other putative influences on child depression, such as parent education (Torvik et al. 2020), seem to be explained by shared genes.

While adoption- and CoT-based parenting studies are valuable, their utility is limited in three ways. First, they focus on how specific measured parent phenotypes relate to child outcomes. This is useful for identifying or disqualifying specific causes. For instance, maternal depression after but not during pregnancy influences offspring emotional problems above genetic confounds (Hannigan et al. 2018; Gjerde et al. 2019). However, it is difficult to know *a priori* how best to identify and measure salient parent traits, and at what point in childhood to measure them. A second limitation is that the direction of effects is often not – or cannot be – assessed. Child-to-parent effects exist (Ahmadzadeh et al. 2019; Eley, Napolitano et al. 2010; McAdams et al. 2015; Narusyte et al. 2008), so where studies do not distinguish these from parent-to-child effects, associations cannot be interpreted as evidence for parental effects on child development. Third, reliance on highly specific samples (e.g. adoptees) may preclude large-scale data collection and reduce the generalisability of findings.

Novel intergenerational genomic designs present opportunities to address these limitations and extend insights from twin and adoption research. The Relatedness Disequilibrium Regression (RDR) method was introduced as a technique for estimating genetic influence without bias from the family environment (Young et al. 2018). A less emphasised attribute of this method is that RDR can estimate environmental effects of parents. RDR measures the effect of parent behaviour indexed by their genome – termed ‘genetic nurture’ (Kong et al. 2018). In RDR, estimates of genetic nurture capture all behaviours of both parents that are influenced by their common genetic variation, and that affect the child’s trait. This is the first advantage of RDR: it removes the reliance on identifying and measuring specific parent traits. Unlike those estimated in adoption and CoT designs, the parent effect is global and atemporal. Genetic nurture indexes any lasting parent effect from any parent phenotype, up to the time-point at which the child’s phenotype was measured. Additional advantages of RDR are that it allows child-versus parent-driven effects to be disentangled from each other and uses scalable population-based family samples rather than adoptive or twin families. A previous study detected genetic nurture for child depression using a similar method (Jami et al. *in press*). However, the analysis was underpowered, maternal and paternal effects were not modelled simultaneously, and mediating parent phenotypes were not explored.

Here, we apply RDR to 11,598 parent offspring trios in the Norwegian Mother Father and Child Cohort Study (MoBa) to index environmental effects of parents on children’s anxiety and depression symptoms using the parental genome. Alongside parental genetic nurture, RDR estimates direct child genetic effects, and the covariance between direct and nurturing effects. We use available maternal trait measures to test whether genetic nurture is mediated by self-reported maternal depression and anxiety. Additionally, we directly compare RDR results against traditional pedigree estimates, using 26,086 pairs of relatives (cousins, siblings, half-siblings and twins) from the child generation of MoBa.

## Methods

### Sample

The Norwegian Mother, Father and Child Cohort Study (MoBa; Magnus et al. 2016) is a prospective population-based pregnancy cohort study conducted by the Norwegian Institute of Public Health. Pregnant women were recruited from across Norway from 1999 to 2008. The women consented to initial participation in 41% of the pregnancies. The total cohort includes 114,500 children, 95,200 mothers and 75,200 fathers. To date, 98,110 individuals who are part of a trio (both parents and a child) from MoBa have been genotyped (Supplementary Table 1). As part of the Intergenerational Transmission of Risk (ITOR) subproject, MoBa has also been linked to Norwegian registry data containing pedigree and twin zygosity information. Version 11 of the quality-assured MoBa data files were used, released in 2018. Written informed consent was obtained from all participants upon recruitment. The establishment and data collection in MoBa was previously based on a license from the Norwegian Data Protection Agency and approval from The Regional Committee for Medical Research Ethics, and it is now based on regulations related to the Norwegian Health Registry Act. The current study was approved by The Regional Committee for Medical Research Ethics.

RDR and pedigree analyses required slightly different subsamples of MoBa families. For RDR, we used genotyped trios with available child anxiety and depression data. Siblings in the child generation were removed so that trios were independent families. For pedigree analyses, we retained all available cousins, siblings, half-siblings and twins in the child generation with anxiety and depression data, without requiring the presence of genotype data. Since only a subset of the families in MoBa were genotyped, all individuals included in genomic analyses were also within the sample used for pedigree analyses.

### Measures

Our two outcome variables are well-validated and reliable quantitative measures of childhood anxiety and depression symptoms at age 8. Anxiety was measured using the 5-item version of the Screen for Child Anxiety Related Disorders (SCARED) (Birmaher et al. 1999). Depression was measured using the 13-item Short Moods and Feelings Questionnaire (SMFQ) (Angold et al. 1995). Both questionnaires were rated by mothers using three-point Likert response scales. Phenotypes were regressed on child sex prior to analyses.

To test whether measured parent emotional phenotypes explained any possible genetic nurture effects, we used the mother-rated Hopkins Symptoms Checklist (SCL-8) (Tambs & Røysamb, 2014). The SCL-8 assesses self-report anxiety and depression symptoms experienced during the last two weeks. To obtain a reliable measure of maternal emotions and behaviours that children are consistently exposed to, we capitalised on the longitudinal data in MoBa and constructed a common factor of SCL-8 scores at five time-points: 15 weeks of pregnancy, 30 weeks of pregnancy, child age 6 months, 18 months, and 8 years. As has been demonstrated with childhood data (Cheesman et al. 2018), a stable factor composed of measurements at multiple time points is more likely than individual scores at a single time point to capture a heritable core trait. Applying this approach to assess maternal symptoms leads to a measure that captures stable exposure to maternal depression symptoms, rather than exposure associated with temporary fluctuations. Mediation of any genetic nurture effect by stable maternal emotional symptoms would be indicated by observed change in the genetic nurture point estimate when using the factor score as a covariate.

### Genotype quality control

The current MoBa genomic dataset comprises imputed genetic data for 98,110 individuals (∼32,000 parent offspring trios), derived from nine batches of participants, who make up four study cohorts. Within each batch, parent and offspring genetic data were quality controlled separately. Quality control exclusion criteria for individuals were: genotyping call rate <95%, or autosomal heterozygosity >4 standard deviations from the sample mean. Quality control exclusion criteria for SNPs (single nucleotide polymorphisms) were: ambiguous (A / T and C - G), genotyping call rate <98%, minor allele frequency <1%, or Hardy-Weinberg equilibrium P-value <1 × 10-6. Population stratification was assessed, using the HapMap phase 3 release 3 as a reference, by principal component analysis using EIGENSTRAT version 6.1.4. Visual inspection identified a homogenous population of European ethnicity and individuals of non-European ethnicity were removed. The parents and offspring datasets were then merged into one dataset per genotyping batch, keeping only the SNPs that passed quality control in both datasets. Phasing was conducted using Shapeit 2 release 837 and the duoHMM approach was used to account for the pedigree structure. Imputation was conducted using the Haplotype reference consortium (HRC) release 1-1 as the genetic reference panel. The Sanger Imputation Server was used to perform the imputation with the Positional Burrows-Wheeler Transform (PBWT). The phasing and imputation were conducted separately for each genotyping batch. Supplementary Table 1 contains details of the numbers of SNPs and individuals in each batch. More detailed information about the cohorts, quality control and imputation can be found at https://github.com/folkehelseinstituttet/mobagen.

We conducted post-imputation quality control, selecting SNPs meeting the follow criteria: high imputation confidence scores (INFO >0.8 on average across batches), minor allele frequency >0.05, Hardy-Weinberg equilibrium p <1×10-6, non-multiallelic, non-duplicated.

Before calculating relatedness matrices for RDR, we removed individuals who were not part of a complete genotyped trio, and pruned down to 451,442 variants in approximate linkage equilibrium using an r2 threshold of 0.5 (to reduce the size of the matrices and therefore the computational burden of the analyses).

After data management, a final sample of 25,828 genotyped parent child trios were identified. Almost 11,600 of these trios also had child emotional symptom data.

### Statistical analyses

#### I. Genomic analysis of trios: Relatedness Disequilibrium Regression (RDR)

The RDR method (Young et al. 2018) allows the estimation of parent and child genetic effects on traits. This is achieved by extending a standard genomic method for estimating heritability – single-component GREML (Genomic-Relatedness based restricted Maximum-Likelihood) (Yang et al. 2011) – to include individuals’ parents. Standard GREML estimates the variance explained by common SNPs by comparing a matrix of pairwise genomic similarity for unrelated individuals across genotyped SNPs to a matrix of their pairwise phenotypic similarity, using a random-effects mixed linear model. Instead of using the random variation in genetic similarity among unrelated individuals, RDR capitalises on the random variation in genetic similarity between parents and offspring, which arises due to random segregation, and is independent of environmental factors.

There are two versions of RDR: one uses identity-by-descent (IBD) relatedness, which distinguishes parts of the genome that are inherited from common ancestors, and the other uses common SNP-based relatedness. We used the SNP version since it has similar properties to the IBD version but has greater statistical power. Rather than estimating a single genetic variance component, RDR estimates three. The first estimates the *direct* effect of children’s own genetic variation on their trait. This is independent of the effect of being reared by biological parents. Importantly, a direct genetic effect is only direct in the sense that it does not stem from another individual’s genotype. Notably, mechanisms by which individuals evoke and select environments based on their genotype, are essential in how genes lead to phenotypes (Plomin, 2014), and these are included in estimates of direct genetic influence.

The second variance component estimates the effect of parent genetics on the child trait, controlling for child genetic effects: ‘genetic nurture’. Any parent genetic effect over and above child-driven direct effects must be an *indirect* genetic effect, where parents’ genes affect child traits by influencing parent behaviours and the rearing environment they provide.

The third component captures variance in the offspring phenotype attributable to covariance between the direct and nurturing genetic effects, known as ‘passive gene-environment correlation’ (see Supplementary Figure 1 for detail on this concept). This somewhat abstract variance component is easier to understand when considering the conditions for the estimate to be zero. Specifically, the direct-nurturing genetic covariance would not explain any phenotypic variation if only one generation contributes genetic effects to the child trait, or if different SNPs contribute to child and parent genetic influences, such that loci have only either direct or indirect effects.

Finally, the residual component captures environmental effects on the trait of interest that are not correlated with measured parent genetic variation, the effects of variants not tagged by genotyped SNPs (e.g. rarer), and measurement error.

In practice, the variance components are estimated by regressing phenotypic resemblance on three genomic relatedness matrices simultaneously. The first is similar to the matrix used in standard GREML: the genome-wide genetic relatedness of the children in the sample. The second and third represent the genetic relatedness of the parents and the genetic covariance between children and parents.

Notably, the genotypes of mothers and fathers are combined to allow estimation of the average effect of *both* parents, rather than only mothers as in a similar model, M-GCTA (Eaves et al. 2014), which estimates the effects of the mother and child genomes and of their covariance. Therefore, before constructing the parent genomic relatedness matrix, maternal and paternal genotypes are summed and standardised to have mean zero and variance two (Young et al. 2018).

All RDR analyses included 10 ancestry principal components and genotyping batch, both derived from the child generation, as covariates. Analyses were performed in the GCTA software using the --reml-no-constrain flag to allow variance components to take positive and negative values (Yang et al. 2011).

To test whether any genetic nurture effect was partially explained by parent emotional symptoms, we re-ran the RDR models adding a measure of stable maternal emotional symptoms as a covariate. We also tested the individual time-specific maternal measures as covariates.

To test whether any of the variance components were biased because some children were genotyped in a different batch to their parents, we re-ran the RDR models using only individuals who were genotyped as a complete trio (90% of the analysis sample).

#### II. Classical pedigree modelling

To compare RDR results to a traditional quantitative genetic (non-genomic) design, we implemented a univariate pedigree model (Neale & Cardon, 1992). As in the classic twin design, this model allows estimation of genetic, shared environmental, and non-shared environmental (residual) influences on anxiety and depression symptoms at age 8. The model used phenotypic correlations among twins, siblings, half-siblings and cousins in the child generation, to derive estimates based on the following specifications: genetic correlations are 1.00, 0.50, 0.25, and 0.125 for identical twins, non-identical twins/siblings, half-siblings and cousins respectively, and shared environmental correlations are 1.00 for all siblings and 0.00 for cousins.

#### Comparison of variance components from RDR and pedigree models

Before comparing RDR and pedigree results, it is essential to note the differences between variance components derived from the models. First, with respect to child genetic effects, the pedigree genetic component denotes a broader effect of any genetic sequence variation, not only common additive effects tagged by measured variants, as in RDR applied to common SNPs. Second, regarding family environmental effects, the shared environment component in the pedigree model is broader than the genetic nurture estimate using RDR. The shared environment captures at least three sources of variance that genetic nurture does not: parent behaviours not tagged by common SNPs, shared environments *beyond* parent behaviours, and effects of any correlation between the shared environment and genetic components (passive gene-environment correlation). Third, whereas pedigree and twin models assume passive gene-environment correlation is absent (i.e. that genetic and environmental effects are uncorrelated), RDR can directly estimate it as the covariance between genetic and environmental (genetic nurturing) effects.

Supplementary Figure 1 and its accompanying text explore overlaps and distinctions between the concepts of genetic nurture, passive gene-environment correlation, and shared environment.

#### Software

Genome-wide relatedness matrices were constructed using python, and RDR models were run using GCTA (Yang et al. 2011). Pedigree analyses were conducted in R using the structural equation modelling package OpenMx (Neale et al. 2016).

## Results

### RDR model

Results from our RDR analyses using genomic data to disentangle child and parent genetic effects on anxiety and depression symptoms at age 8 are shown in Figure 2. Direct effects of children’s own common genetic variation (yellow bar) explained 5% of the variance in anxiety symptoms (se=0.07), and 19% (se=0.07) of the variance in depressive symptoms. Genetic nurture (dark green bars) had a negligible influence on variation in child anxiety, but explained 14% (se=0.07) of the variance in child depression. For depression, the estimate of the phenotypic variance explained by covariance between direct and indirect genetic effects (i.e. passive gene-environment correlation) was negative (−16%; se=0.07; pale green bars).

**Figure 2.**
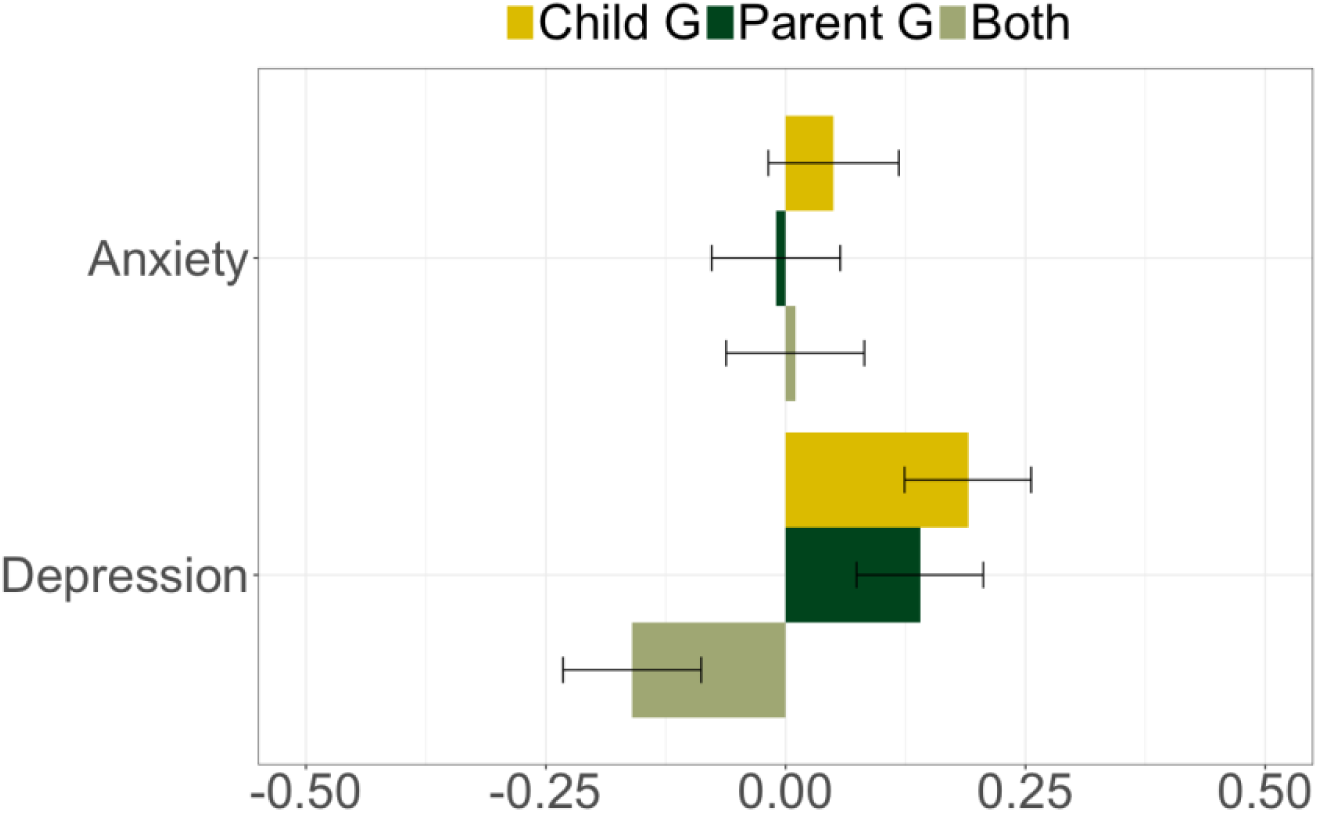
Variance components from the RDR model. Labels= variance explained by direct child genetic effects (yellow), parent genetic effects (i.e. genetic nurture; dark green), and by covariance between the direct and nurturing genetic effects (‘Both’; pale green). Bars=standard errors. Not shown: residual variance affecting the phenotypes, including parent genetic effects not tagged by common SNPs, individual-specific environmental effects, chance factors and error.

See Supplementary Table 2 for full RDR results including sensitivity analyses exploring mediators, batch effects, and the effect of constraining components to take positive values. In the sensitivity analysis exploring mediation, maternal emotional symptoms partially explained the genetic nurture effect on childhood depression. This manifested as the genetic nurture effect, but not the other variance components, being substantially attenuated when including a covariate capturing stable maternal anxiety and depression symptoms. The genetic nurture effect on child depression dropped from 14% to 5%. Most individual time-specific maternal measures also led to attenuation of the genetic nurture effect when included as covariates, even some that were measured during pregnancy.

Our results were robust to any effects of children being genotyped in a different batch to their parents. Variance components were unchanged after restricting the sample to trios who were genotyped together.

Supplementary Figure 2 visualises the RDR results in a path diagram, with estimates of paths for direct and nurturing effects, and of the correlation between them.

### Pedigree model

Our key finding of genetic nurture for child depression but not anxiety was supported by our pedigree analysis (Figure 3). Shared environmental influences contributed 13% (se = 0.05) of the variation in depression, but we observed no robust evidence of shared environmental effects on anxiety. The pedigree heritability estimates were similar for anxiety and depression (∼37%). This contrasts to the pattern of heritability estimates from RDR, in which child genetic effects explained more variance in depression than anxiety (19% versus 5%, respectively). See Supplementary Figure 3 for a path diagram version of the pedigree results.

**Figure 3.**
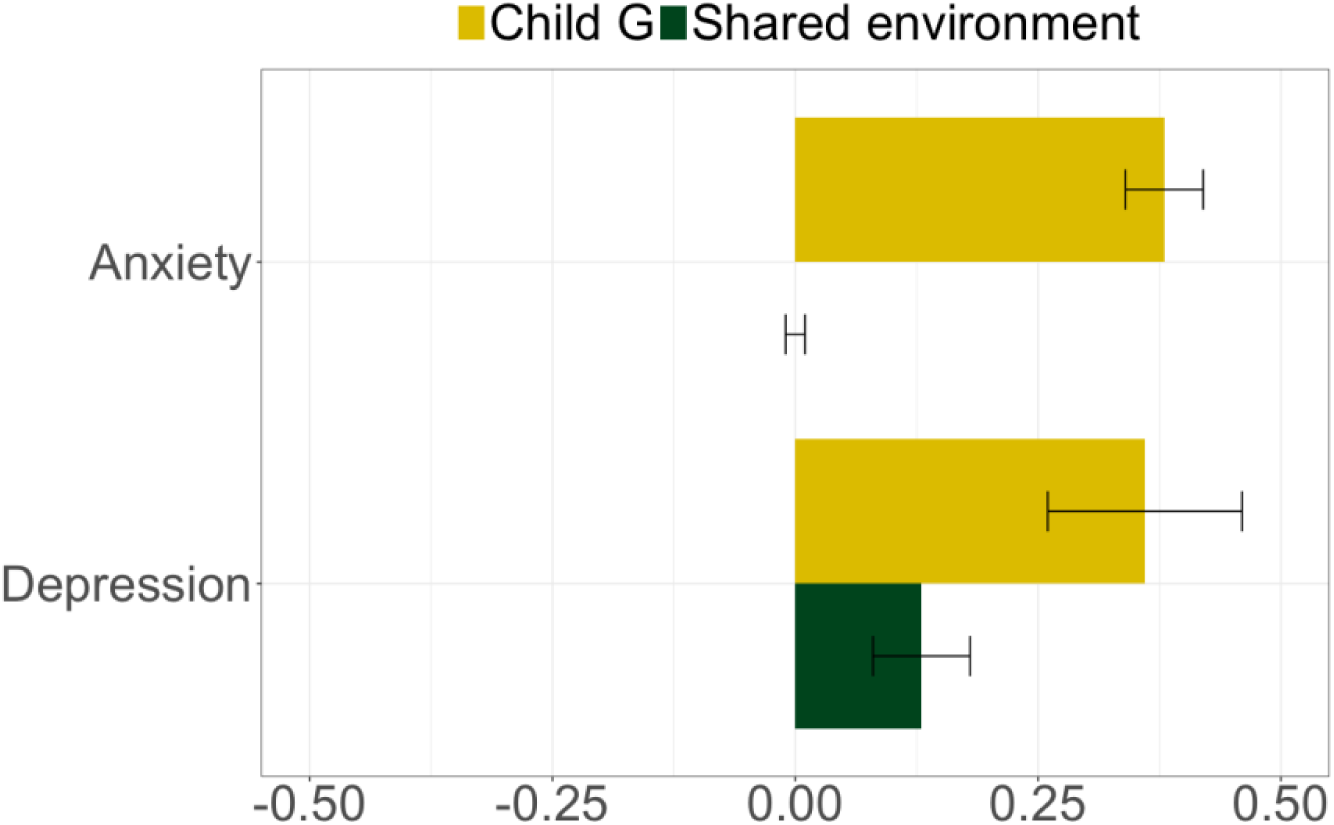
Pedigree variance components. Labels= variance explained by genetic effects (i..e pedigree heritability; yellow), variance explained by environmental influences that increase resemblance among siblings in the same family (dark green). Bars = standard errors. Not shown: Residual variance, which includes non-shared environmental effects and error. The effect of any covariance between genetic and shared environmental influences is not estimated in this model.

## Discussion

We used genomic data from parent-offspring trios to estimate indirect parent genetic effects (genetic nurture) separately from direct child genetic effects on anxiety and depression symptoms at age 8. Results from the RDR model suggested that depressive symptoms were significantly influenced by both genetic nurture and direct genetic effects. Genetic nurture is an environmentally mediated effect of genetic origin, indexing behaviours of both parents that are influenced by their common genetic variation and that influence child depression, over and above direct child genetic effects. We found that the genetic nurture effect was partially mediated via an observed measure of maternal anxiety and depression, which explained 64% of the original genetic nurture effect (14%). In contrast, individual differences in anxiety symptoms were not significantly influenced by parental genetic nurture, and direct effects of common genetic variants explained just 5% of the phenotypic variance.

### Genetic nurture for child depression symptoms

Our genomic evidence of an environmental effect of parents on child depression, but not anxiety, was supported by pedigree-based evidence for shared environmental influence on depression, but not anxiety. This aligns well with findings from epidemiological and twin studies. Unlike anxiety, depressive symptoms are uncommon in pre-pubertal children, but when they do occur, are typically a response to psychosocial risks, such as perinatal issues, abuse, and bullying (Jaffee et al. 2002; Maughan et al. 2013). Twin data also suggest a greater role for family environmental factors in child depression than anxiety. In one twin study, shared environmental effects contributed 18% of the variance in depressive symptoms in middle childhood, but none for anxiety (Cheesman et al. 2018). The use of RDR adds to our knowledge of parent effects from pedigree methods, because genetic nurture is simultaneously specific to parents and global. In contrast, estimates of parental influence from twin studies (i.e. the shared environment) are not specific to parents, and those from the children of twins (CoT) method rely on phenotypic measurement of specific parent traits.

### Mediation by maternal anxiety and depression

Our analysis adjusting for parent emotional symptoms suggested that maternal anxiety and depression mediate the genetic nurture effect on childhood depression. This is compatible with adoption- and CoT-based evidence for phenotypic effects of maternal depression (Natsuaki et al. 2014). This environmental influence could reflect a learning mechanism directly related to maternal emotional symptoms, or could be mediated through secondary parenting-related behaviours such as disorganized home environments (Rapee et al. 2009; Natsuaki et al. 2014).

### Negative covariance between direct and nurturing genetic effects on child depression

This negative component suggests that genetic variation that is associated with higher depression risk when present in children, has an opposing effect (*reducing* child depression risk) when present in the parents. On the face of it, this result is difficult to explain and we suggest that it should be interpreted with caution. Notably, negative covariance components have been reported in previous RDR analyses of multiple phenotypes including creatinine levels and age of first child in men (Young et al. 2018), and in a Maternal-GCTA analysis of gestational weight gain (Warrington et al. 2018). If our finding is real, it could be explained by maternal emotional sensitivity. In other words, genes that lead mothers to identify depressive symptoms in their children could be associated with parent behaviours that reduce those same symptoms.

### Pedigree and genomic heritabilities

For child anxiety, the pedigree heritability estimate was moderate, despite the small genomic signal from RDR. This is compatible with findings from other samples (Cheesman et al. 2017). However, it is surprising that the SNP-based effects are lower for anxiety than depression, given that they are similar traits, with similar pedigree heritability estimates. One possible explanation is that anxiety, more than depression, is influenced by gene-by-shared environment interactions. These interactions may be captured in pedigree heritability (genes make identical twins more similar than non-identical twins; shared environmental factors make both sets of twins more similar, so the interaction may further increase identical twin similarity) but not RDR heritability estimates, since the latter does not compare children who share a family environment.

### Limitations

We acknowledge potential sources of bias affecting the RDR results. First, although supportive of the notion of parental influence on child depressive symptoms, the genetic nurture estimate may also capture residual population stratification, assortative mating, and indirect genetic effects from siblings.

Second, the genetic nurture effect, and the mediation effect, could be partly generated by the use of maternal ratings of child outcomes. Maternal reports of child depression symptoms may reflect their own emotional symptoms. As a result, an apparent parental genetic nurture effect may in fact be a direct genetic effect on parents’ own traits. This phenomenon has been evidenced in developmental twin studies, in which rater bias may cause shared environmental variance to be overestimated (Bartels et al. 2004). However, if rater bias explained the environmental effect for depression, then we might expect to see some evidence for this with anxiety too. The lack of a genetic nurture effect for anxiety in our RDR and pedigree models suggests that maternal report is not entirely reflective of maternal phenotype. Measurement issues will be clarified with the upcoming availability of child, teacher, and clinician reports in MoBa.

Third, selective participation, genotyping and attrition might have reduced coverage of families experiencing more severe emotional symptoms. Depression-linked attrition has been demonstrated in the Norwegian Twin Panel (Tambs et al. 2009). Stronger parent effects might occur at the tails of the distribution of child emotional problems. Future linkage of MoBa with Norwegian registry data will help to investigate and control for participation biases affecting the genotyped trios.

### Future directions

Future studies could seek to identify behaviours other than maternal emotional symptoms that explain why the parent genome independently influences child depression. Since common genetic variants only capture a subset of the genetic component of complex traits involved in parenting, our observed genetic nurture effect on childhood depression is likely to be larger than we estimate here. This implies substantial scope for identifying heritable parent phenotypes with effects on child depression symptoms. However, individual parenting phenotypes may have small effect sizes, and are usually only modestly heritable (∼20%)(Klahr & Burt 2014). Researchers should therefore continue to explore wider social factors that might affect parenting, and in turn, childhood anxiety and depression.

Mediators of genetic nurture might depend on socioeconomic context. For example, poverty increases the magnitude of the effect of maternal depression on diverse child outcomes (Goodman et al. 2011). In the future, RDR could be adapted to allow moderation effects to be tested.

The RDR method could be broadened to address multivariate and longitudinal questions, as has been done with twin designs (Boomsma et al. 2002). For example, does genetic nurture persistently influence depressive symptoms across development? Twin research suggests that the effects of parents (and passive gene-environment correlation) will be stronger earlier in life (Hannigan et al. 2017; Rice, 2010; Thapar & McGuffin, 1994). This is partly because with increasing age less time is spent in the family environment, and individuals have greater capacity to choose their experiences and express their genetic predispositions. Genomic tools could be used to strengthen and develop findings from traditional designs.

### Summary

Our genomic approach strengthens the evidence base regarding parent effects on child emotional development, by directly accounting for shared genes, and by indexing the child’s environment using the parental genome. Quantifying the contribution of parents is crucial for the understanding of how anxiety and depression develop in childhood, and potentially how to target modifiable causes. Our results are supportive of parent effects on depressive symptoms at age 8. The genetic nurture effect on depressive symptoms was partially mediated by maternal anxiety and depression. Though the effect size is modest, this suggests that efforts to alleviate maternal emotional problems could help to prevent child depression.

## Data Availability

MoBa data can be accessed through an application process detailed here: https://www.fhi.no/en/studies/moba/for-forskere-artikler/research-and-data-access/.

## Acknowledgements

The work was supported by the Norwegian Research Council (Grant Number 262177). TE is part-funded by a program grant from the UK Medical Research Council (MR/M021475/1), and by the National Institute for Health Research (NIHR), the Biomedical Research Centre at South London, Maudsley NHS Foundation Trust and King’s College London. EY, and TAM are supported by the Norwegian Research Council (grant number 288083). AH and LJH are supported by the South-Eastern Norway Regional Health Authority (grant numbers 2018058 and 2018059). The views expressed are those of the author(s) and not necessarily those of the NHS, the NIHR or the Department of Health. TAM and YIA are supported by a Sir Henry Dale Fellowship jointly funded by the Wellcome Trust and the Royal Society to TAM (Grant Number 107706/Z/15/Z). RC is supported by an ESRC studentship. The Norwegian Mother, Father and Child Cohort Study is supported by the Norwegian Ministry of Health and Care Services and the Ministry of Education and Research. We are grateful to all the participating families in Norway who take part in this on-going cohort study. We thank the Norwegian Institute of Public Health (NIPH) for generating high-quality genomic data as part of the HARVEST collaboration, supported by the Research Council of Norway (#229624). We also thank the NORMENT Centre for providing genotype data, funded by the Research Council of Norway (#223273), South East Norway Health Authority and KG Jebsen Stiftelsen. We further thank the Center for Diabetes Research, the University of Bergen for providing genotype data and performing quality control and imputation of the data. PRN was funded by the ERC AdG SELECTionPREDISPOSED ((#293574), the Stiftelsen Kristian Gerhard Jebsen, the Trond Mohn Foundation, the Norwegian Research Council (#240413/F20), the Novo Nordisk Foundation (#54741), the University of Bergen, and the Western Norway health Authorities (Helse Vest; PERSON-MED-DIA and #911745). OAA is funded by the Research Council of Norway (#223273 NORMENT, #248778 BIOBANK, #273291 ResilieMent), the South-East Norway Regional Health Authority (2019-112), and the European Union’s Horizon2020 Research and Innovation Action Grant (#847776 CoMorMent), and the KG Jebsen Stiftelsen.

